# Defining the determinants of under-vaccination in migrant populations in Europe to improve routine and COVID-19 vaccine uptake: a systematic review

**DOI:** 10.1101/2021.11.08.21266058

**Authors:** Alison F Crawshaw, Yasmin Farah, Anna Deal, Kieran Rustage, Sally E Hayward, Jessica Carter, Felicity Knights, Lucy P Goldsmith, Ines Campos-Matos, Fatima Wurie, Azeem Majeed, Helen Bedford, Alice S Forster, Sally Hargreaves

## Abstract

Diverse migrant populations in Europe are at risk of under-immunisation and have recently shown lower levels of COVID-19 vaccination intent and uptake. Understanding the determinants of vaccine uptake in migrants is critical to address immediate COVID-19 vaccination inequities, and longer-term will help improve coverage for routine vaccinations, aligning with the goals of the new Immunisation Agenda 2030. We did a systematic review following PRISMA guidelines and using a PICOS framework (PROSPERO CRD42020219214; MEDLINE, CINAHL, PsycINFO databases, 1 January 2000 – 14 September 2021) exploring barriers and facilitators to vaccine uptake and determinants of under-vaccination in migrants in the EU/EEA, UK, and Switzerland. We categorised barriers/facilitators using the ‘5As’ Determinants of Vaccine Uptake Taxonomy. 5259 data sources were screened, with 67 studies included from 16 countries, representing 366,529 migrants. Access barriers were most commonly reported (language, literacy and communication barriers; practical and legal barriers to accessing/delivering vaccination services; service barriers, including lack of specific guidelines and knowledge of healthcare professionals) for key vaccines including MMR, DTP, HPV, influenza, polio, COVID-19 vaccines. Acceptance barriers were mostly reported in Eastern European and Muslim communities for HPV, measles, and influenza vaccines. We identified 23 determinants of under-vaccination in migrants, including geographical origin (where 25/26 (96%) studies showed significance) – particularly African/Eastern European origin; recent migration; being a refugee/asylum seeker; higher income; parental education level; no healthcare contact in the past year; and lower language skills. Facilitators of migrants’ vaccine uptake included tailored vaccination messaging, community outreach and ‘nudging’ interventions. Migrants’ barriers to accessing healthcare are already well documented, and this review confirms their role in limiting vaccine uptake. These data hold immediate relevance to strengthening vaccination programmes in high-income countries, including for COVID-19. Our findings suggested that targeted, evidence-informed strategies are needed to address access and acceptance barriers to vaccination in migrants, including the development of migrant-sensitive and adaptable vaccination services and systems, unambiguous public health messaging, and coproduction of tailored interventions.

## Introduction

Some migrant populations are known to be at risk of under-immunisation (1-3) and have been involved in recent outbreaks of vaccine-preventable diseases in the EU/EEA. (4) The severe health inequities exposed by the COVID-19 pandemic (5-8), including barriers to accessing vaccination services (9), have highlighted the need for novel strategies to improve engagement with under-immunised groups, address barriers to COVID-19 vaccine uptake, and facilitate countries in meeting their vaccination targets, relieving their health systems and reopening their economies. (9, 10) Evidence shows lower COVID-19 vaccine uptake in some migrant and ethnic minority populations, despite these groups being some of the worst affected by the disease. (5) Adolescent and adult migrants may be particularly at risk of under-immunisation for routine vaccinations and excluded from initiatives to promote catch-up vaccination on arrival in some European countries. (11) Migrants also face well-documented barriers to accessing healthcare (2, 12) but it is unclear to what extent this impacts on their ability to access vaccination services or how cultural, personal and language barriers also influence vaccine uptake. (9) Despite known gaps in uptake, there is limited research exploring these factors and how levels of vaccination coverage and uptake vary within and between migrant sub-populations.

Understanding the factors that influence low vaccination coverage and uptake in migrants, and identifying which sub-populations of migrants are affected, is critical to driving improvements in vaccination programmes and national vaccination strategies, including in the immediate term for COVID-19. It also supports key objectives of the World Health Assembly’s new Immunisation Agenda 2030 (IA2030) (13) to improve vaccine coverage for vaccine-preventable diseases and improve equitable access for all populations. (14) At present, inconsistent use of terminology and a range of theoretical models complicate the discourse around vaccination (and migrant health more generally) and may contribute to the design of interventions which fail to account for the full range of reasons for sub-optimal vaccination (15, 16). The ‘5 As’ taxonomy (17) captures most determinants of vaccine uptake and categorises them according to access (ability to reach or be reached by recommended vaccines), affordability (financial and non-financial costs), awareness (of need/availability of recommended vaccines), acceptance (the degree to which individuals accept, question, or refuse vaccination) and activation (the degree to which individuals are nudged towards vaccination) (Table 2). There is an urgent need to investigate the relative contributions of these various factors to sub-optimal vaccine uptake in migrant populations to inform the development of evidence-based interventions to improve vaccine equity. We therefore did a systematic review to identify 1) barriers and facilitators to vaccine uptake in migrants (categorised using the 5As) and 2) determinants of under-vaccination, to improve uptake and coverage of routine, catch-up and COVID-19 vaccination in diverse migrant populations in the EU/EEA.

## Methods

We did a systematic review according to PRISMA guidelines (18) and registered with the International Prospective Register of Systematic Reviews (PROSPERO; CRD42020219214).

### Inclusion and exclusion criteria

We included primary research studies that included data on barriers or facilitators (primary outcome) to vaccine uptake or determinants of under-vaccination (secondary outcome) in migrant populations living in one of 30 EU/EEA countries, the UK and Switzerland, published between 2000-2021 in any language. Studies involving healthcare professionals (HCPs) working with migrant populations were included to capture provider- and system-level perspectives pertaining to our primary outcome. Migrants were defined as foreign-born (or, in the case of children, had at least one migrant parent); a barrier was defined as a factor that hindered vaccine uptake while a facilitator was defined as a factor that supported or promoted it. A determinant of under-vaccination was defined as a factor statistically associated (p<0·05) with incomplete coverage or uptake of recommended vaccines, or where uptake/coverage was statistically significantly lower compared with the reference population. We included all vaccines in this analysis, including COVID-19 vaccines. The inclusion and exclusion criteria, developed using a PICOS framework (19), are outlined in full in Table 1. Studies were excluded if they did not contain data from one of the listed countries, were published outside the specified date range, contained non-disaggregated migrant population data, did not meet the key definitions, or were non-primary research articles.

**Table 1.** Inclusion and exclusion criteria, using PICOS framework. (19)

|  | **Inclusion criteria** | **Exclusion criteria** |
| --- | --- | --- |
| **Population** | - Adult, adolescent, and child migrants (foreign-born) and children of migrants (under 16 years of age, with at least one migrant parent) residing in the UK, Switzerland or one of 30 EU/EEA countries*<br>- HCPs (doctors, nurses, healthcare assistants, etc) who work with/have worked with the above populations | - Migrant status not defined by country/region of birth or not defined<br>- Data not disaggregated between migrants and non-migrants<br>- Data not collected from one of the listed countries |
| **Intervention** | - Vaccination | n/a |
| **Control** | - No comparator or control was selected for this review | n/a |
| **Outcome** | - Barriers and facilitators to vaccine uptake in migrant populations (primary outcome)<br>- Determinants of under-vaccination in migrant populations (secondary outcome) | n/a |
| **Study design** | - Primary research | - Non-original research articles (e.g. reviews, commentaries, editorials, case reports and guidelines on vaccination) |
| **Other** | - Published in any language | - Did not meet definitions for primary or secondary outcomes<br>- Papers reporting immunity gained through natural disease (as opposed to vaccination) |

\*Austria, Belgium, Bulgaria, Cyprus, Croatia, Czech Republic, Denmark, Estonia, Finland, France, Germany, Greece, Hungary, Iceland, Ireland, Italy, Latvia, Liechtenstein, Lithuania, Luxembourg, Malta, Netherlands, Norway, Poland, Portugal, Romania, Slovakia, Slovenia, Spain, Sweden. N/A not applicable.

### Search strategy

MEDLINE, CINAHL and PsycINFO databases were searched for primary research in any language published between 1 January 2000 and 14 September 2021, combining free text terms and subject headings relating to (migration) AND (vaccination) AND (determinants) (see Tables S3-S4). Grey literature sources and bibliographies of included studies were also hand-searched. Pre-2000 studies were excluded to keep findings relevant to recent migrant population flows, policy and events; literature on COVID-19 was included. Records were imported into EndNote, and duplicates deleted. Title/abstract and full-text screening were independently carried out by two reviewers (AFC and JC/KR/AD) using Rayyan QCRI. (20) Papers not written in languages spoken by the research team (English, Spanish, French) were translated using Google Translate. The selection process is shown in Figure 1.

**Figure 1.**
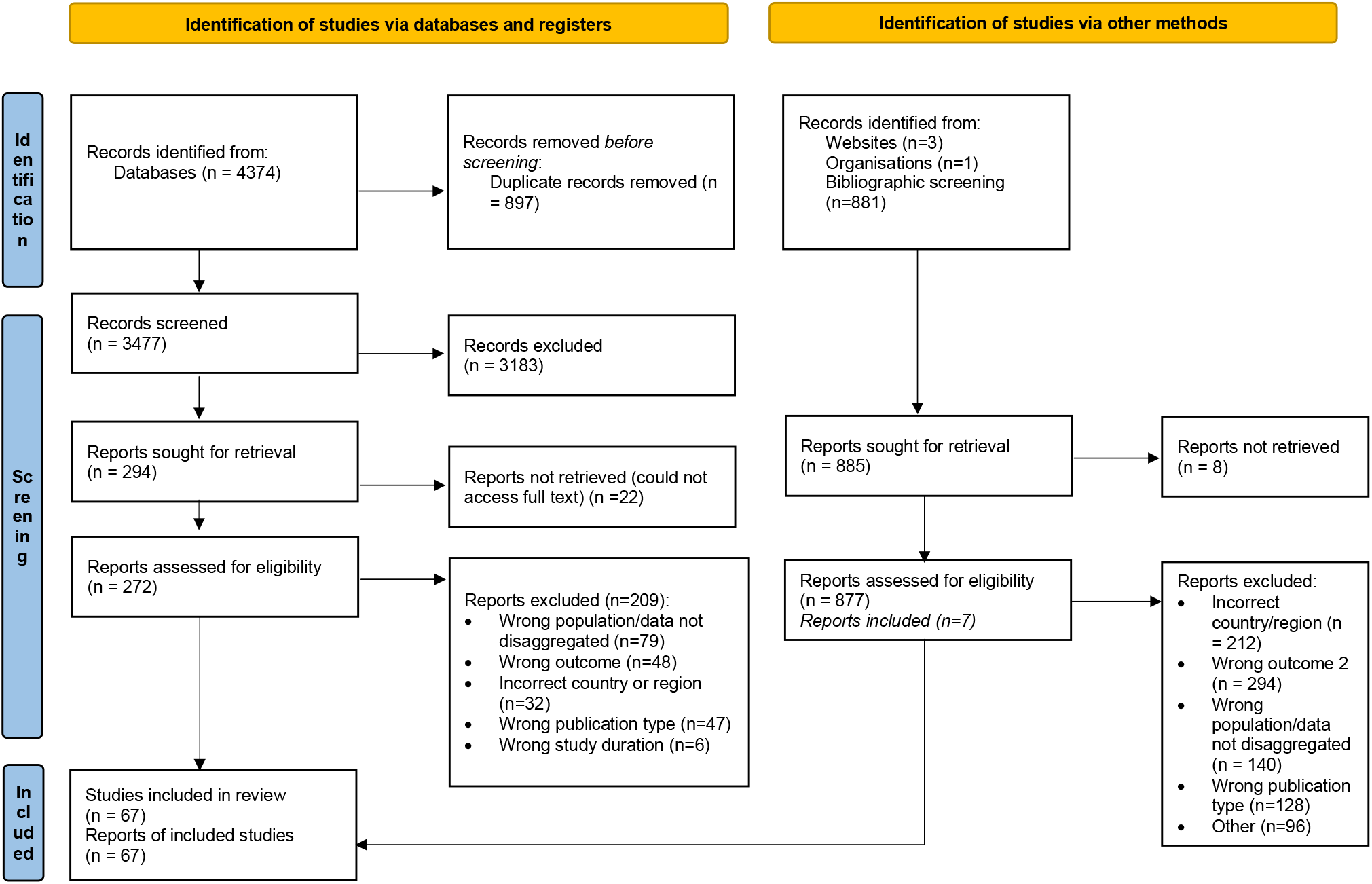
Study selection process, as shown by PRISMA 2020 flow diagram for new systematic reviews which included searches of databases, registers, and other sources. Reproduced from: (18)

### Data extraction

Data were independently extracted by two reviewers (AFC and YF) using a customised form (developed and piloted for the review), including location and year of study, study design, methodology, population/sub-population, sample size, vaccine(s), vaccination type (e.g., routine childhood immunisations), determinants and rates of under-vaccination, barriers and facilitators to vaccine uptake and reasons for vaccination decisions. Discrepancies at any stage were resolved by consensus.

### Quality assessment

Quality of all included studies were independently assessed by two reviewers (AFC and YF) using JBI Critical Appraisal Tools (21), with parameters of low (<49%), medium (50-79%) and high (80-100%) study quality. Data were not excluded based on study quality, but this information informed the narrative synthesis and discussion.

### Data synthesis and analysis

Extracted data were tabulated and results presented as reported in the studies. All data were synthesised narratively. Qualitative data were first analysed thematically to identify factors influencing uptake, then categorised using the ‘5 As’ Taxonomy for the Determinants of Vaccine Uptake (17) (Table 2), and further classified by emergent sub-themes. Data synthesis and analysis were carried out by two reviewers (AFC and YF) in consultation. Studies that explored HCPs’ awareness of migrant health, guidelines, and policy in relation to vaccine uptake were classified as access, rather than awareness, factors.

**Table 2.** The ‘5 As’ Taxonomy for the Determinants of Vaccine Uptake. Reproduced from: (17)

| **Domain** | **Definition** |
| --- | --- |
| Access | The ability of individuals to be reached by, or to reach, recommended vaccines |
| Affordability | The ability of individuals to afford vaccination, both in terms of financial and non-financial costs (e.g., time) |
| Awareness | The degree to which individuals have knowledge of the need for, and availability of, recommended vaccines and their objective benefits and risks |
| Acceptance | The degree to which individuals accept, question, or refuse vaccination |
| Activation | The degree to which individuals are nudged towards vaccination uptake |

## Results

5259 data sources were screened (title/abstract, n=4362; full-text, n=1149), of which 67 studies were included in the systematic review (primary outcome, n=43; secondary outcome, n=37); 42 focused on ‘foreign-born’ migrants (not otherwise defined) or children of migrants, while the remainder focused on asylum seekers and refugees (n=10), undocumented migrants (n=3), homeless migrant children (n=1), European Roma (n=2) and HCPs who had worked with migrants (n=8) (papers containing multiple population groups were counted more than once). The included studies had a combined sample size of 366,529 migrants and 641 health professionals. Most studies reported on measles-containing vaccines (n=18), human papillomavirus (HPV) vaccine (n=17), or diphtheria, tetanus, or pertussis-containing vaccines (n=16); two studies looked at COVID-19 vaccination in migrants. Studies were conducted in 16 countries; and were cross-sectional (n=36), cohort (n=12), case-control (1), qualitative (n=16) or other (n=2) in design. 67/70 papers were quality appraised (3 study designs did not have an appropriate checklist), with a mean score of 82% (range: 22-100%). Detailed characteristics of included studies are shown in Supplementary Table S1.

### Barriers and facilitators to vaccine uptake in migrant populations

Forty-three studies (7, 22-63) addressed the primary outcome. Access and acceptance were the most common themes, with awareness, affordability and activation reported to a lesser extent. Unique sub-themes relating to barriers (n=20) and facilitators (n=18) to uptake were defined and are summarised in Table 3 (further details are shown in Tables S5-S6).

**Table 3.** What are the barriers and facilitators to vaccine uptake in migrants?

| Barriers | Facilitators |
| --- | --- |
| **Access** |  |
| • **Language, literacy, and communication barriers** (23, 25, 33-36, 40, 41, 43, 50, 52, 55-57, 61, 62, 64)<br>• **Resource and capacity constraints** (36, 38, 44, 49, 55, 58, 62)<br>• **Practical barriers** (31, 36, 38, 39, 41, 42, 44, 56)<br>• **Legal barriers** (39, 42, 66)<br>• **Distrust of health system/authorities, sense of alienation and disempowerment** (7, 31, 36, 55-57)<br>• **Specific provider-level barriers** (31, 39, 43, 45, 55, 57-59) e.g., health professionals lack specific knowledge of migrant entitlements, catch-up vaccination guidelines; missed opportunities to vaccinate | • **Social integration** (31, 36, 41, 45, 61), e.g., engaging with health or vaccination system, having citizenship<br>• **Service coordination, organisation, and infrastructure** (24, 44, 55)<br>• **Culturally competent and migrant-sensitive care** (26, 31, 35, 38, 50, 56, 57, 60) e.g., inclusive services and policies, alternative access points<br>• **Tailored information sources** (7, 50, 56)<br>• **Vaccination policy** (60), e.g. policy to vaccinate in absence of vaccination card<br>• **Trust in the provider/system/State** (31, 57) |
| **Affordability** |  |
| • **Direct costs** (25, 29, 39, 55)<br>• **Indirect costs** (56) e.g., cost of travelling to vaccination appointment<br>• **Competing priorities** (31, 55, 56) | • **Cost offsetting** (29, 33, 36, 41, 45, 56) e.g., free vaccination, insurance cover<br>• **Convenience** (35, 55) e.g., walk-in clinics rather than pre-booked appointments; flexible appointments |
| **Awareness** |  |
| • **Lack knowledge about disease/need for vaccination** (7, 25, 27, 32, 34, 36, 39, 40, 42, 43, 48, 51, 52, 55, 57, 61)<br>• **Lack knowledge of entitlement to vaccination** (36, 42)<br>• **Personal health stewardship** (42, 63) e.g., knowing own medical/vaccination history<br>• **Misinformation or lack of information** (7, 48, 55-57) e.g., about the vaccine or its availability | • **Health promotion/awareness** (32, 42) e.g., health educational programmes, being aware of benefits of vaccination |
| **Acceptance** |  |
| • **Worries about vaccine safety/side effects** (27, 31-36, 51, 53, 55, 56, 61)<br>• **Cultural, religious, and social barriers** (33, 34, 46, 51-53, 61), e.g., stigma around specific vaccines, vaccination unfashionable in home country<br>• **Distrust of health system/authorities, sense of alienation and disempowerment** (7, 31, 52, 55, 56)<br>• **Misinformation or lack of information** (7, 27, 33, 53, 56, 57)<br>• **Low perception of risk of disease or importance of vaccination** (7, 27, 34, 39, 55-57, 61)<br>• **Vaccination not physician-recommended** (27) | • **Positive perceptions of vaccination** (31, 32, 34, 35, 64)<br>• **Positive social norms** (31, 33, 35, 36, 57) , e.g., normalisation of vaccination<br>• **Tailored approaches, information, and messaging** (35, 52), e.g., emphasising that HPV vaccine prevents cervical cancer, rather than a sexually-transmitted infection<br>• **Access to credible information sources** (34, 48, 53)<br>• **Provider recommendation** (27, 32, 48, 51, 53, 61) |
| **Activation** |  |
| • **Lack of information/practical support from HCPs when desired** (61)<br>• **Blanket approaches** (55) e.g., vaccination reminders sent via letter/text message not suitable for transient Roma populations | • **Catch-up vaccination initiatives** (44, 47) e.g., on-arrival health screening and vaccination for asylum seekers, mass vaccination campaigns<br>• **Mandates** (51), e.g. mandatory workplace vaccination<br>• **Provider recommendation** (51)<br>• **Health promotion and education** (39)<br>• **Culturally tailored and community-based interventions** (36, 44, 54, 55) e.g., face-to-face communication, personalised reminders, community advocates |
| **Other** |  |
| • **Lack vaccination documentation/record** (38, 42, 62, 63) |  |

#### Access to vaccination

##### i. Language, literacy, and communication barriers

Low literacy (36, 50, 55), language barriers and lack of interpreting services (23, 25, 33, 34, 36, 40, 41, 43, 50, 52, 55, 62, 64, 65) were widely reported barriers to uptake. Specific barriers for European Roma were highlighted, with providers unaware of the difference between Roma and Romanian and lack of access to Romani-speaking interpreters. (36, 55) Health professionals reported that the time allocated for vaccination appointments (10-15 minutes) was unrealistic when faced with communication barriers. (55) These barriers were mirrored at the system level in the lack of accessible, tailored or translated information about vaccination, available in different formats, for migrant populations. (23, 25, 33, 35, 41, 43, 50, 52, 55-57, 61, 65) For example, some Moroccan, Turkish and Somali populations said they valued oral information more and felt the written format of the information they received was not appropriate. (35, 52) In the absence of translated or accessible information, many migrants reported that they turned to alternative and unregulated sources, such as Google, social media, friends, and family. (22, 33, 53, 56, 57)

##### ii. Practical and legal barriers to accessing healthcare and services

Practical and legal barriers to accessing healthcare (whether perceived or real) and vaccination delivery models, meant that many migrants experienced exclusion and challenges accessing vaccination services. Insecure housing and frequent change of (or no fixed) address, due to destitution, undocumented status, or the dispersal process for new migrants made attending vaccination appointments and follow-ups more difficult. (31, 38, 41) Jackson and colleagues (36) in 2017 highlighted the digital exclusion felt by European Roma who lacked access to the internet; while vaccines delivered through mainstream channels were not accessible for certain sub-populations. (31, 36, 65) Fears of being charged for care or asked about immigration status, distrust of HCPs and authorities based on rumours or past experiences of discrimination, difficulties registering with a GP, or being refused care, were also highlighted. (7, 31, 36, 55-57, 65) These fears were a major barrier to COVID-19 vaccination access in two UK studies of asylum seekers, refugees, and migrants with precarious immigration status, who expressed concern that they would be de-prioritised or excluded from the COVID-19 vaccine roll-out because of their status. (7, 56) Migrants (n=10) interviewed after government announcements to widen COVID-19 vaccine access to undocumented migrants also remained unaware that they could be accessed free of charge and without immigration checks. (56)

##### iii. Knowledge of migrant vaccination among healthcare professionals (HCPs)

Poor HCP knowledge of migrants’ entitlements to healthcare and vaccination guidelines (e.g., vaccination of individuals with incomplete/uncertain immunisation status) was highlighted, with instances of patients being wrongly refused access to primary care or not offered recommended catch-up vaccinations. (31, 45, 57, 62) A French study of GPs found wide variation in vaccination practices for newly-arrived migrants with no vaccination record, with some GPs reluctant to revaccinate those with uncertain immunisation status due to concerns about reactogenicity. (58) There were also missed opportunities to vaccinate; Moura et al. (2019) found that, in a Portuguese study of access to tetanus vaccination, two thirds of migrants without a tetanus vaccination in the previous ten years had visited a family/general physician in the past 12 months. (45)

##### iv. Vaccination policy and guidelines

The absence of policies promoting opportunities for vaccination was also a major barrier in some settings. This included lack of guidelines on offering BCG vaccinations against tuberculosis to migrants in most surveyed European countries (66) and not assessing the immunisation status of refugees on arrival in Hungary. (42) Louka et al. (2019) (39) found that only 36·1% of participating asylum seekers and refugees were asked about their vaccination status by official authorities, healthcare workers or non-governmental organisations (NGOs) on arrival in Greece or the Netherlands.

##### v. Local variability in approach and coordination

Mellou et al. (2019) (44) discussed the challenge of implementing vaccination campaigns in camps across Greece, with activities mostly carried out by NGOs, and determined by factors such as hosted population size and site resources. Higher vaccination coverage at larger camps was attributed to more organised, frequent, and effective vaccination campaigns compared to in smaller camps. A Welsh study reported variability in local procedures and resource allocation between asylum dispersal sites, including differences in accepting verbal history as proof of vaccination status, staff allocation, and follow-up procedures. (49)

##### vi. Resource and capacity constraints

Staff shortages, including of bi-lingual HCPs, interpreters (36) and cultural mediators (44), were barriers, particularly in camps (44) and reception settings. (49, 62) In two studies, the mode of determining vaccination status and need for catch-up vaccination through mapping activities in the absence of a record was deemed too resource intensive. (38, 62)

##### vii. Timing and engagement with services

Timing of vaccination during the migration or asylum-seeking process was highlighted. One study suggested that low participation in a vaccination programme in Hungary was due to it being a transit, rather than a destination, country. (42) However, the opposite effect was also observed (lower uptake at the destination), which the authors attributed to a switch in priorities of asylum seekers upon reaching their destination. (39) It was separately hypothesised that lower coverage in Syrian refugees in Greek camps could be due to higher turnover of this population relative to others, due to a more straightforward asylum process. (44)

##### viii. Facilitators to vaccine uptake: Cultural competence, integration, and engagement, and alternative access points

Cultural competence of HCPs and migrant-sensitive services and policies facilitated uptake. (26, 31, 38, 50, 60) In Sweden, for example, all newly-arrived migrant children are invited to attend a health dialogue meeting with the school nurse, which helps to establish trust early and determine health and vaccination needs. (38) A UK study increased immunisation uptake in unaccompanied asylum-seeking children by training and supporting social workers, primary care staff and young people and promoting the importance of vaccination and regular monitoring. (26)

Social integration and engagement with the healthcare system also had a positive association with uptake. Two studies found that migrants (adults and homeless children) who had been in contact with the healthcare system or a GP in the previous year had a significantly lower risk of being insufficiently immunised. (41, 45) Similarly, regular contact with the local health services, (31) closer geographic proximity to a GP, (36) and greater integration into the local community and familiarity with health services, (61) were associated with greater engagement and vaccination uptake.

Migrants in a UK study valued convenience of access and preferred familiar settings requiring minimal travel. They suggested that access points in the community, such as walk-in clinics at food banks, community centres and charities, could help improve COVID-19 vaccine uptake, as well as encouraging and facilitating migrants to register with primary care. (56)

#### Acceptance of vaccination

##### i. Social norms, cultural acceptability, and stigma as barriers

Four studies reported barriers stemming from cultural acceptability and stigma around specific vaccines such as HPV. (33, 51-53) For example, Somali Muslim communities felt HPV vaccination promoted promiscuous sexual behaviour and was unnecessary as Somali women are expected to not engage in pre-marital sex. Religious and personal reasons were also more often cited as a reason for refusing tetanus vaccination among foreign-born construction workers compared to Italians. (51) Two studies suggested that negative social norms and different recommendations around vaccination in migrants’ countries of origin were a barrier, with vaccination considered “unfashionable” in Poland, and not recommended in pregnancy. (33, 53)

##### ii. Concerns about safety and side effects

Worries about “overloading” the immune system with multiple or combined childhood vaccines, and side effects including death, paralysis, or the potential effects on an unborn child, in the case of specific vaccines such as HPV and MMR were highlighted by a range of migrant groups. (27, 31-34, 36, 51, 53, 56, 61) Studies also suggested that migrants’ vaccination perceptions (including anti-vaccination sentiment) were influenced by a reliance on information and messages from their home countries, including friends, family, media, social media, and other online resources. (33, 53) Vaccine anxieties around the MMR/autism controversy were also reported to be a barrier to the uptake of other vaccines. (53) Concerns about the contents and potential side effects of the newly introduced COVID-19 vaccines were raised in a UK study, where three quarters of the migrants interviewed (23/32; 72%) were reluctant to accept a COVID-19 vaccination and stated they would need more information before making a decision. (56) In a study of undocumented migrants, concerns about side effects were less about life and death, and more about the implications for accessing healthcare. Here, the primary concern about vaccinating their children was that side effects might require medical attention and thus contact with the health system, which they were keen to avoid. (31)

##### iii. Low perceived value, risk of vaccine-preventable diseases, or importance of vaccination

In nine studies (7, 27, 33, 34, 39, 53, 56, 57, 61), uptake was hindered by a belief that vaccination was unimportant or not fully protective, or because patients felt they lacked credible information about the need for vaccination. Some Romanian and Romanian Roma parents considered contracting measles a rite of passage for their child and a way to build natural immunity against the disease, or considered them unnecessary or ineffective (particularly influenza and MMR vaccines). (55) Five studies highlighted how this lack of information could lead to exposure to misinformation from unofficial sources, presenting further barriers to uptake. (7, 33, 53, 56, 57) Some migrants explained their reluctance to vaccinate against COVID-19 was because they felt it was not needed, and they preferred to rely on natural remedies, their immune system or self-isolation to prevent infection. (7, 56)

##### iv. Alienation and disempowerment

Distrust of the healthcare system and fear of being questioned about one’s legal status was reported as a barrier both to accessing, and accepting, routine and COVID-19 vaccination. (7, 31, 52, 55, 56, 65) Interestingly, Ukrainian migrants in Poland expressed distrust of their home country’s vaccination policy, public servants, and medical system, but this made them more accepting of vaccination in Poland, where they felt the quality of vaccines and the healthcare system were higher. (57)

##### v. Facilitators relating to acceptance

Decisions to vaccinate were influenced by a general positive attitude towards vaccination and its benefits, confidence in the advice of HCPs, positive religious beliefs about vaccination, and normalisation of vaccination. (27, 31, 32, 34-36, 64) Re-framing the language and messaging around vaccination was shown to be effective in addressing cultural barriers; for example, emphasising that HPV vaccination prevents cervical cancer, rather than a sexually transmitted infection (52), and linking the benefits of vaccination to religious teachings (e.g., that vaccination can help maintain good health). (35) Having access to a ‘trusted information source’, often from a medical background, was also important (48, 53) and HCP recommendations carried weight. (27, 32, 48, 51, 53, 61)

#### Awareness of need for, and availability of, vaccination

Knowledge-related barriers in migrants included low health literacy or knowledge of the need for vaccination or boosters (25, 27, 32, 36, 39, 40, 42, 48, 51, 61), lacking knowledge about the disease, or its relationship to the vaccine (particularly in the case of cervical cancer and HPV vaccine) (7, 25, 34, 48, 52, 56, 57), unfamiliarity with the immunisation schedule or need for boosters, or where to access them (43, 48, 51, 55, 63), and lacking knowledge or evidence of one’s own vaccination history. (38, 42, 62, 63) Many migrants commented that they struggled to find credible and trustworthy information about vaccination in their own language. Two studies found that migrant adolescents had limited knowledge about the existence of common vaccines, including measles and polio (27), and were unlikely to actively seek out vaccine-related information. (61) Few studies measured the effect of knowledge on vaccine uptake; although one study suggested health educational programmes at reception centres could help asylum seekers integrate into host societies. (42)

#### Affordability of vaccination (financial and non-financial)

##### i. Financial affordability: barriers and facilitators

Few studies investigated financial barriers; however, cost was found to be prohibitive when assessed hypothetically (25, 39) or where self-payment was required. (29) Romanian and Polish participants in two UK studies reported that the cost of vaccines in their country of origin was a barrier. (33, 36) Newly-arrived migrants also highlighted the indirect costs associated with getting a vaccine, such as the cost of travel to appointments (56), and lack of clarity around payment for health services. (55)

Free-of-charge vaccination, or having private health insurance, facilitated uptake. (29, 33, 36, 45) The introduction of a cost-free HPV programme in Denmark was associated with an 11-fold increase in uptake percentage among migrants. (29) Migrants with precarious immigration status said that if they could be confident there would be no associated costs, more of their community would present for COVID-19 vaccination. (56)

##### ii. Non-financial affordability: barriers and facilitators

In a study of undocumented migrants in Sweden, although parents had a positive attitude to vaccination or intended to vaccinate, attending vaccination appointments for their children became a decreasing priority as their child got older, due to competing priorities and uncertain social situations. (31) Competing priorities were also a barrier for Romanian and Romanian Roma migrants and health professionals reflected that pre-booked appointments were poorly attended by this population, whereas offering walk-in vaccination clinics improved attendance. (55)

#### Activation and nudging towards vaccination

Few studies explored how factors relating to activation could influence uptake. One study identified a lack of practical support, exchanges, and recommendations from HCPs around vaccination, when they were desired, as a barrier to uptake. (61)

In terms of facilitators, face-to-face communication and outreach (e.g., during community visits) were generally effective and well received by the Romanian and Roma communities (36, 55), and helped to gain their trust. Personalised vaccination reminders had a larger positive effect on the uptake of childhood vaccines in non-Western mothers compared to Danish mothers. (54) Initiatives that built trust and shared responsibility through local partnerships and collaboration were also effective. (24, 44, 55) Health professionals suggested that, although costly, involving community members as vaccine advocates could help promote vaccination in communities that had experiences measles outbreaks. (55)

### Determinants of under-vaccination in migrant populations

Thirty-seven studies (23, 24, 28-30, 41, 42, 44, 45, 47, 49, 51, 59, 67-90) addressed this secondary outcome. We identified 23 specific determinants of under-vaccination in migrant populations (geographical origin; recent migration; lower acculturation; gender or sex; age; being a refugee/asylum seeker; income; healthcare contact; health insurance; housing insecurity; region of residence; dispersal site; smaller refugee camp; not having citizenship; comorbidity; being in an influenza risk group; and seven parental characteristics, including: younger maternal age; education level; language difficulties; unemployment; one or both parents born overseas; first generation children; larger family size). Geographical origin and recent migration were the factors most associated with under-vaccination. Determinants of under-vaccination are summarised in Table 4 (for further details, see Table S2). Only adjusted analyses were reported, where relevant. Where studies were conducted with a mixed population (migrant and non-migrant), only variables that could be attributed with certainty to the migrant population (e.g. geographical origin) were extracted. Only determinants where a statistical association was found were reported.

**Table 4.**
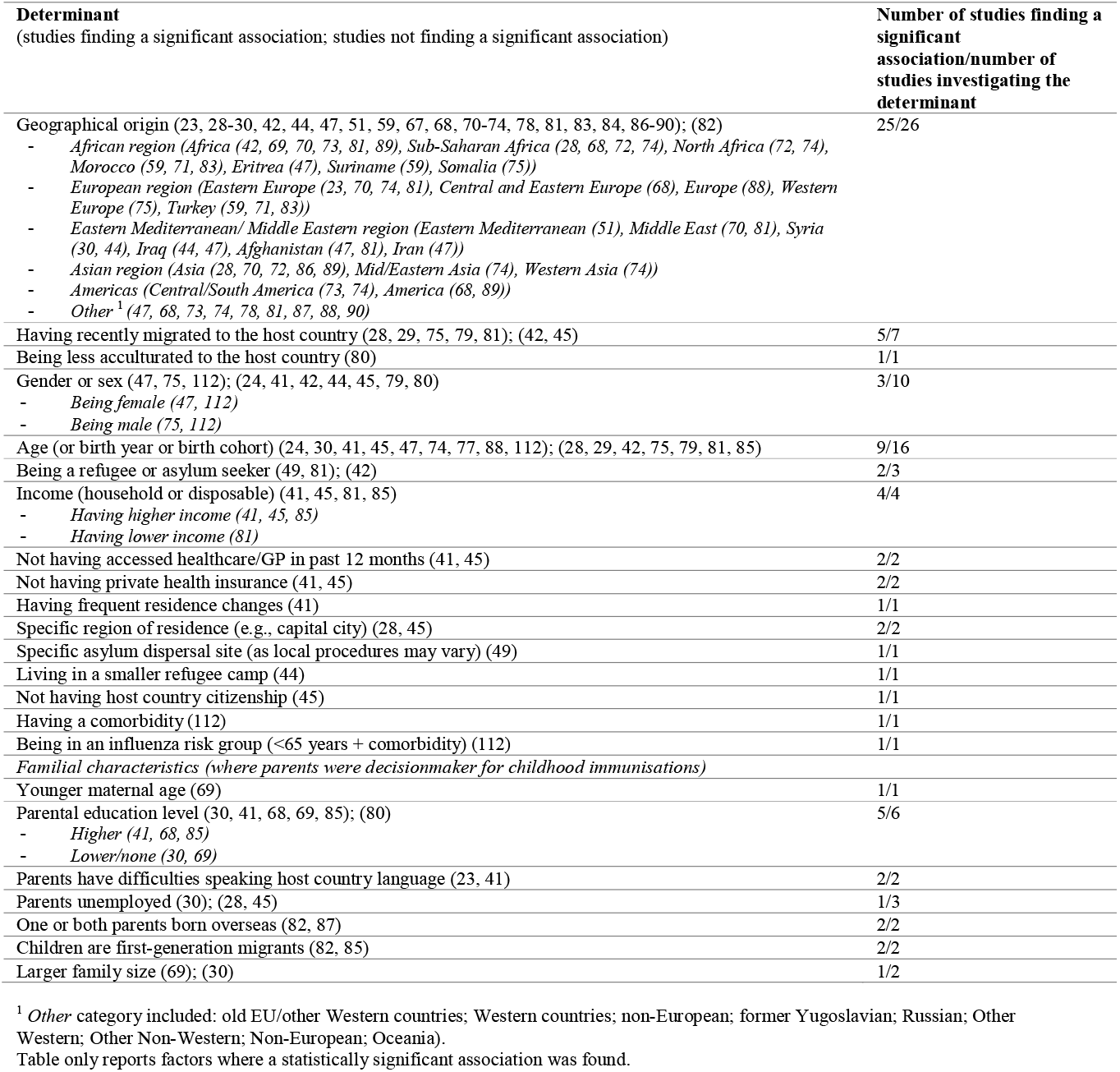
Determinants of under-vaccination in migrants. Lists are shown where relevant to illustrate the direction of the association and related citations (geographical origin; gender; income; education level).

Twenty-five (23, 28-30, 42, 44, 47, 51, 59, 67, 68, 70-74, 78, 81, 83, 84, 86-90) out of 26 studies found a statistically significant (p<0.05) association between geographical origin and under-vaccination. Of these, 16 studies found an association specifically with African origin (Africa (42, 69, 70, 73, 81, 89); sub-Saharan Africa (28, 68, 72, 74); North Africa (72, 74); Morocco (59, 71, 83); Eritrea (47); Suriname (59); Somalia (75)). Five studies found a specific association with Eastern/Central European origin (23, 68, 70, 74, 81); three studies with Turkish origin (59, 71, 83); six studies with Eastern Mediterranean/Middle Eastern origin (Eastern Mediterranean (51); Middle East (70, 81); Syria (30, 44); Iraq (44, 47); Afghanistan (47, 81); Iran (47)); six studies with Asian origin (28, 70, 72, 74, 86, 89), and two studies with Central/South America origin (73, 74) (See Table 4 for full list of associations).

Six (28, 29, 75, 79-81) out of eight studies found that under-vaccination was significantly associated with more recent migration to the host country (28, 29, 75, 79, 81) or lower acculturation with the host society (80). Other common predictors of under-vaccination included higher income (3 out of 4 studies; (41, 45, 85), being a refugee/asylum seeker (2/3 studies; (49, 81)), having not accessed healthcare in more than a year (2/2; (41, 45)), having no private health insurance (2/2; (41, 45)) and region of residence (2/2; (28, 45)). Specific familial characteristics, such as parental education level, difficulties speaking the host country language or larger family size were also associated with under-vaccination. Two studies (41, 85) found that higher income and higher parental education level were associated with under-vaccination of HPV vaccine in children.

We did not identify a strong overall association with gender/sex or age in the data. Seven (24, 41, 42, 44, 45, 79, 80) out of 10 studies found no association with gender/sex and seven (28, 29, 42, 75, 79, 81, 85) out of 16 studies found no association with age. Of the nine studies reporting an association with age, the direction of association varied across samples, and the broadly different reference groups mean it is not possible to draw reliable conclusions. One study found asylum-seeking girls were more likely to be under-vaccinated than asylum-seeking boys, (47) while another study found that Somali boys were more likely to be un-vaccinated for MMR than Somali girls (75); this effect was moderated by age, with a faster decline in MMR vaccination coverage rates over time for Somali boys than girls, indicating some level of intentionality in MMR refusal which the authors suggested may be related to a fear of autism, a condition more prevalent in boys, in the Somali community. (75)

## Discussion

We have reported data on barriers and facilitators to vaccine uptake and defined key determinants of under-immunisation in migrant populations, summarising data on 366,529 migrants living in EU/EEA countries, the UK, and Switzerland. These data hold immediate relevance to strengthening vaccination programmes in high-income countries, including for COVID-19. Access barriers were very common in the literature and related to language, literacy and communication barriers, practical and legal barriers to vaccination services and systems, and service barriers (including lack of dedicated resourcing, specific guidelines, and training/knowledge of healthcare professionals) for key vaccines, including MMR, DTP, HPV, influenza, polio, COVID-19 vaccine. Acceptance barriers were mostly reported in Eastern European and Muslim communities, for HPV, measles, and influenza vaccines, suggesting they may be localised to certain populations, vaccines, and contexts. We identified 23 specific determinants of under-vaccination in migrant populations; particularly prominent, and showing significant associations in published studies, were geographical origin and recent migration, with migrants from the African Region and Eastern European Region most likely to be under-vaccinated, a finding which has immediate policy and planning implications. However, a range of other determinants were identified suggesting that the reasons for under-vaccination of migrants are highly variable and influenced by context.

Among the limited number of studies reporting facilitators to vaccine uptake, tailored vaccination messaging (based on specific perceptions, beliefs and/or barriers), community outreach and ‘nudging’ interventions (e.g., personalised reminders) were shown to be effective. Our findings suggested that targeted, evidence-informed strategies are needed to address key access and acceptance barriers to vaccination in migrants, including migrant-sensitive and adaptable services and systems, provision of clear and accessible health information, and interventions designed with specific migrant communities and tailored to their needs.

The breadth of vaccine access barriers – practical, legal, and administrative – experienced by migrants and identified in this review were significant. Migrants’ barriers to accessing healthcare are already well documented (91-94), and this review confirms their role in limiting vaccine uptake. This included barriers from gaps in healthcare provider knowledge around catch-up vaccination, an area where experts have called for more guidelines. (10, 11) At the policy level, national vaccination strategies and guidelines vary considerably across Europe and many countries do not specifically include or target refugees and migrants in their plans. (12, 95) Recent steps have been taken in Europe to widen access to COVID-19 vaccination for undocumented migrants and marginalised populations following recommendations by international and EU bodies (96-99), including through more accessible distribution points and reducing entitlement and charging barriers, although migrants’ awareness of these policies or willingness to come forward may be limited. (56, 100) Similar steps should be taken to reduce legal barriers to, and increase opportunities for, migrants to access routine and catch-up vaccination. In the short term, strengthening the capacity of host country health systems to enable more opportunities and novel access points for catch-up vaccination of migrants, particularly adults, may help overcome immunity gaps. Longer term measures should focus on improving coordination of policies, guidelines and vaccination delivery for migrants and mobile populations across European borders, as recently recommended by European policymakers. (101)

In addition to improving intra-regional capacity to monitor and deliver vaccination services to migrants, measures must tackle the systemic barriers to accessing vaccination by creating more culturally competent health systems. Migrants described how they lacked trust in the health system, and struggled to communicate with HCPs, access or understand information about vaccination, and how this led them to avoid care, delay vaccination or turn to alternative sources of information, including social media. HCPs highlighted the additional burden that communication barriers and lack of interpreters imposed on their limited consultation time. Wider research shows that such patient-provider obstacles can result in delayed engagement with, and difficulty navigating, health services and patients being less able to communicate concerns, advocate for themselves and obtain better care. (102-104) This may partially explain why more recently arrived and less acculturated migrants were at greater risk of under-vaccination. The Council of Europe urges that “access to vaccination services should be tailored to the needs of persons in vulnerable situations having difficulties in accessing health services” (105), and our findings demonstrate that migrants need more linguistically, socially, religiously and culturally tailored information, in a variety of formats, to make informed decisions about their health, including vaccination, particularly those who may already be reluctant or hesitant to vaccinate. (106) Producing these types of resources should be prioritised by public health bodies.

The importance of clear public health messaging around vaccination was also highlighted in this review, with examples of misinformation and lack of official information influencing vaccination perceptions and decision-making. A particular challenge during the COVID-19 pandemic has been the need for quick and clear communication during a rapidly evolving situation, much of which has been conducted by politicians rather than public health professionals. Recent evidence shows that “vague, reassuring communication” which is more typical of politicians, who are motivated by short-term goals, does not increase vaccine acceptance and leads to both lower trust and higher endorsement of conspiracy theories. (107) It is possible that for migrant populations facing language barriers, these negative trade-offs are even more pronounced. Therefore, governments should recognise the importance of clear and transparent communication in any vaccination campaign, and after vaccine development continue to invest funds in developing strong communication and vaccine roll-out strategies to gain and maintain the trust of – and reach – their entire population. Existing research evidence around effective vaccine communication, and new toolkits to combat vaccine misinformation produced during the pandemic, provide useful guidance. (108-110)

This review has some limitations. Included studies came from only 16 of the 32 review countries; therefore, this review is not fully representative of the European region and largely focuses on Western Europe, highlighting the urgent need for more data on vaccination uptake disaggregating by migrant status in European countries, which is rarely collected by national data systems. The lack of uptake data for the COVID-19 vaccine in diverse migrant populations has been previously highlighted and has undoubtedly hindered evidence-based service delivery. (97, 111) Certain sub-populations and nationalities of migrants were not well reported (e.g., undocumented migrants), and migrant status was generally not well defined or reported in the data, making it difficult to draw firm conclusions.

This review has shown that access to and acceptance of vaccination are key factors influencing vaccine uptake in migrant populations in the EU/EEA. To address these barriers and ensure we meet regional and global targets for immunisation coverage and uptake, multi-level action is needed. Vaccination services should be designed to better meet patients’ social, cultural, and linguistic needs, through the translation and tailoring of information, provision of interpreters, training of HCPs in migrant health/vaccination guidelines and implementation of interventions which facilitate access to vaccination. Tailored and evidence-informed strategies should be co-designed with migrant populations to address specific barriers and perceptions towards vaccines and vaccination in context. Effective and unambiguous communication of public health messages, delivered by trusted messengers, will be vitally important to reach and gain the trust of migrant populations, and to combat the spread of misinformation, as highlighted by the COVID-19 vaccine roll-out. The findings of this review have immediate implications for strengthening national and regional routine immunisation programmes and public health responses to the COVID-19 pandemic.

## Data Availability

All data produced in the systematic review are contained in the manuscript and supplementary files.

## Contributions

AFC and SH had the idea for this review and designed the protocol. ASF, HB, KR, JC, AD, SEH, YF reviewed and commented on the protocol. AFC and YF independently conducted and verified the searches, screening, data extraction and analysis, with validation and support from KR, JC, AD and SEH. AFC and YF wrote a first draft of the paper with input from SH and ASF. All authors discussed the findings and contributed to the review and editing of the final manuscript.

## Acknowledgments

We thank the members of our NIHR Patient and Public Involvement Project Advisory Board: Babatunde Tikare, Larysa Agbaso, Monika Hartmann, Saliha Majeed, Yusuf Ciftci.

## Declaration of Interests

This study was funded by the National Institute for Health Research (NIHR200072) and supported by the ESCMID Study Group for Infections in Travellers and Migrants (ESGITM). AFC, LPG and SH are funded by the NIHR (NIHR300072); AFC and SH are funded by the Academy of Medical Sciences (SBF005\1111). AD and SEH are funded by the Medical Research Council (MRC/N013638/1). JC is funded by an NIHR-in practice clinical fellowship (NIHR300290). KR is funded by the Rosetrees Trust (M775). AM is supported by the NIHR Applied Research Collaboration NW London. The funders did not have any direct role in the writing or decision to submit this manuscript for publication. HB is a member of the National Institute for Health and Care Excellence committee developing guidance on vaccine uptake in the general population. SH is a freelance Senior Editor for *The Lancet Infectious Diseases* and other *Lancet* journals. All other authors declare no competing interests. The views expressed are those of the author(s) and not necessarily those of the NHS, Department of Health and Social Care, or the NIHR. All authors had full access to the full data in the study and accept responsibility to submit for publication.

## Notes

### Competing Interest Statement

The authors have declared no competing interest.

### Clinical Protocols

https://www.crd.york.ac.uk/prospero/display_record.php?RecordID=219214

