## Supplementary Materials for "Defining the determinants of under-vaccination in migrant populations in Europe to improve routine and COVID-19 vaccine uptake: a systematic review"

**Table of Contents**

[**Table S 1. General characteristics of all included studies, including study quality (based on JBI critical appraisal tools and further organised into L (Low: <49%), M (Medium: 50-79%) and H (High: 80%-100%)). Study relevance: a score of direct or indirect denoting the study’s relevance to this review’s aims; N/A = Not applicable or not specified in the study; HPV = Human papillomavirus vaccine, MMR(V) = measles, mumps, rubella (and varicella) vaccine; DTP= diphtheria, tetanus, pertussis vaccine, DTaP= diphtheria, tetanus, acellular pertussis vaccine, IPV = inactivated polio vaccine, OPV = oral polio vaccine, Td/IPV = tetanus, diphtheria, polio vaccine, Hib = Haemophilus influenzae type b vaccine, MenC = Meningococcal group C vaccine, MenAWCY = Meningococcal group A, C, W-135, and Y vaccine, BCG = Bacillus Calmette-Guerin vaccine, COVID-19= coronavirus disease 2019 vaccine.**	2](#_Toc85102408)

[**Table S 2. Statistically significant (p<0.05) determinants of under-vaccination in migrants and details, presented by citation, study location and population.**	17](#_Toc85102409)

**Table S 1. General characteristics of all included studies, including study quality (based on JBI critical appraisal tools and further organised into L (Low: <49%), M (Medium: 50-79%) and H (High: 80%-100%)). Study relevance: a score of direct or indirect denoting the study’s relevance to this review’s aims; N/A = Not applicable or not specified in the study; HPV = Human papillomavirus vaccine, MMR(V) = measles, mumps, rubella (and varicella) vaccine; DTP= diphtheria, tetanus, pertussis vaccine, DTaP= diphtheria, tetanus, acellular pertussis vaccine, IPV = inactivated polio vaccine, OPV = oral polio vaccine, Td/IPV = tetanus, diphtheria, polio vaccine, Hib = Haemophilus influenzae type b vaccine, MenC = Meningococcal group C vaccine, MenAWCY = Meningococcal group A, C, W-135, and Y vaccine, BCG = Bacillus Calmette-Guerin vaccine, COVID-19= coronavirus disease 2019 vaccine.**

| **Study Author(s)** | **Year(s) of study** | **Country** | **Study design** | **Method** | **Vaccine** | **Sample size** | | | **Study population** | **Objective addressed** | **Primary** | **Secondary** | **Study relevance** | **Quality** | |
| --- | --- | --- | --- | --- | --- | --- | --- | --- | --- | --- | --- | --- | --- | --- | --- |
|  |  |  |  |  |  | **Total** | **Of which, migrants & descendants** | |  |  | **Factors identified (5As)** | **Exposure (bold: associated w/ under-immunisation)** |  |  |  |
|  |  |  |  |  |  |  | **(n)** | **(%)** |  |  |  |  |  |  |  |
| Amdisen et al. (1) | 2013-2017 | Denmark | Cohort | Register-based nationwide cohort study | HPV | 161,528 | 16,960 | 10.3%  (1999-2000)  10.7%  (2001-2003) | Danish girls born between 1999 and 2003 | Secondary | n/a | **Geographical origin** | Indirect | 73% | M |
| Bielecki et al. 2020 (2) | 2018 | England, UK | Cross-sectional | Survey | Nasal influenza | Pilot: 2694  Control: 1913  Questionnaire: 128 | Pilot: 1125  Control: 404  Questionnaire; 128 |  | Polish school pupils in Edinburgh and their parents in 2016, 2017 and 2018 | Primary | Access, Acceptance | n/a | Direct | 63% | M |
| Borras et al. (3) | 2003 - 2004 | Spain | Cross-sectional | Telephone Survey | Childhood immunisations: DTaP, OPV, Hib, MenC, MMR | 630 | 60 |  | Native and immigrant children living in Catalonia | Secondary | n/a | **Maternal age, number of children, maternal education, paternal education** | Direct | 63% | M |
| Bouhamam et al. (4) | 2009 – 2010 | France | Cross-sectional | Survey | DTP, BCG, MMR, Pneumococcal conjugate vaccine (PCV), Hepatitis B | 375 | 143 |  | Children aged 9 months to 7 years visiting two public paediatric emergency departments in Marseille | Both | Access | **Geographical origin (mother), parents’ level of spoken French** | Indirect | 63% | M |
| Brockmann et al. (5) | 2014 - 2015 | Germany | Case-control | Analysis of a vaccination Initiative | DTaP/IPV, MMR | 2256 | 2256 |  | Asylum seekers in reception centres | Both | Activation | **Age**, sex, immunisation concept (intervention vs control) | Direct | N/A | |
| Chadenier et al. (6) | 2008 – 2009 | Italy | Cross-sectional | Questionnaire | HPV | 475 | 44 |  | Mothers of eligible daughters attending the practice in 2 health districts in Italy | Primary | Access, Awareness, Affordability | n/a | Indirect | 88% | H |
| Chauhan and Ward (7) | 2016 - 2017 | England, UK | Project evaluation | Questionnaires and in-depth Interviews | Not specified | 14 | 14 |  | Unaccompanied asylum-seeking children | Primary | Access | n/a | Direct | N/A | |
| Cleary et al. (8) | 2009 - 2010 | Ireland | Cohort | Historical cohort study | Influenza | 6838 | N/A | | Mothers of singleton deliveries at University Hospital, Dublin | Secondary | n/a | **Geographical origin** | Indirect | 90% | H |
| De Oliveira Bressane Lima et al. (9) | 2018 - 2020 | Netherlands | Cohort | Evaluation of a mass vaccination campaign | Men ACWY | 154, 757 | 18,844 |  | Adolescents born between January 2001 and December 2005 | Secondary | n/a | **Geographical origin (parents)** | Indirect | 75% | M |
| Devroey et al. (10) | 2009 | Belgium | Cross-sectional | Survey | Not specified | 186 | 88 |  | Students aged 14-17 in the last 4 years of secondary school and in technical classes | Primary | Awareness, Acceptance | n/a | Direct | 89% | H |
| Fabiani et al. (11) | 2011 – 2015 | Italy | Cross-sectional | Analysis of Surveillance data | Rubella | 44,234 | 3,140 |  | Mothers living in Italy | Both | Access | **Geographical** **origin, length of stay, area of residence** | Direct | 88% | H |
| Fernandez de Casadevante et al. (12) | 2012 - 2013 | Denmark | Cohort | Nationwide retrospective cohort study | HPV | 230,032 | 47,790 | 20.80% | Female Danish citizen born 1985-1992 | Both | Affordability | **Geographical** **origin, years lived in Denmark**, year of birth, marital status, area of residence | Direct | 82% | H |
| Fozouni et al. (13) | 2016 | Germany | Cross-sectional | Interviews and analysis of medical records | Polio, measles, tetanus | 229 | 229 |  | Children of refugee families living in German refugee camps | Both | Access | **Age, Geographical** **origin, paternal unemployment,** maternal unemployment, **parental education**, number of siblings, born outside hospital | Direct | 78% | M |
| Freund et al. (14) | 2009 - 2010 | France | Cohort | Prospective cohort study | Influenza (Pandemic 2009/A H1N1) | 882 | 225 |  | Pregnant women >18 years old between 12 – 35 weeks’ gestation | Secondary | n/a | **Geographical origin** | Indirect | 100% | H |
| Godoy-Ramirez et al. (15) | 2013 | Sweden | Qualitative | Semi-structured interviews | Not specified | 10 | 7 |  | Undocumented migrants’ families and nurses | Primary | Access, Affordability, Acceptance, | n/a | Direct | 80% | H |
| Gorman et al. (16) | 2018 | Scotland, UK | Qualitative | Interviews | HPV, Influenza, not specified | 13 | 13 |  | Polish migrant mothers or grandmothers | Primary | Access, Affordability, Acceptance | n/a | Direct | 90% | H |
| Gorman et al., (17) | 2018 | Scotland, UK | Cross-sectional | Questionnaire | Influenza, not specified | 128 | 128 |  | Parents/guardians of Polish students in Edinburgh | Primary | Acceptance, Awareness | n/a | Direct | 44% | L |
| Grandahl et al. (18) | 2011 | Sweden | Qualitative | Focus group discussions | HPV | 50 | 50 |  | Immigrant women learning Swedish | Primary | Acceptance, Awareness | n/a | Direct | 90% | H |
| Gutierrez Hernando et al. (19) | 2003 | Spain | Cross-sectional | Analysis of medical records | Not specified | 73 | 73 |  | Immigrant children aged between 3 months and 14 years | Primary | Awareness | n/a | Direct | 38% | L |
| Hansen et al. (20) | 2009 - 2011 | Norway | Cross-sectional | Register-based nationwide cross-sectional study | HPV | 90,842 | 25,082 |  | Mothers and fathers of teenage girls resident in Norway | Secondary | n/a | **Geographical** **origin** **(father)** | Indirect | 100% | H |
| Harmsen et al. (21) | 2012 | Netherlands | Qualitative | Focus group discussions | Childhood immunisations/not specified | 33 | 33 |  | Immigrant mothers | Primary | Acceptance, Awareness, Access | n/a | Direct | 80% | H |
| Hertzum-Larsen et al. (22) | 2009 - 2015 | Denmark | Cohort | Register-based nationwide cohort study | HPV | 260,251 | 26,539 |  | Girls born 1996 - 2003 | Secondary | n/a | **Geographical** **origin, birth year** | Direct | 91% | H |
| Jackson et al. (23) | 2013 - 2014 | UK | Cross-sectional | Mix of individual and group interviews | Not specified | 174 | 61 |  | Travelling and European Roma communities | Primary | Awareness, Acceptance, Access, Affordability, Activation | n/a | Indirect | 90% | H |
| Jiménez-García et al. 2006 (24) | 2003 | Spain | Cross-sectional | Questionnaire | Influenza | 27,791 | 863 |  | National sample of Spain’s non-institutionalized population (aged ≥ 6 months) | Secondary | n/a | **Age** | Indirect | 56% | M |
| Jimenez-Garcia et al. 2008 (25) | 2004 - 2005 | Spain | Cross-sectional | Database analysis | Influenza | 908 | 908 | 12.4% | Sample of adult subjects (ages 16 years and over) living in Madrid | Secondary | n/a | **Age, sex, comorbidity, risk group,** healthcare worker (yes/no). | Direct | 88% | H |
| Klok-Nentjes et al. (26) | 2016 | Netherlands | Cross-sectional | Questionnaire | Childhood immunisations/not specified | 320 | 320 |  | Undocumented migrants and their children living in the Amsterdam region | Primary | Access | n/a | Indirect | 22% | L |
| Laenen et al. (27) | 2013 - 2014 | Belgium | Cross-sectional | Survey | Influenza, Pertussis | 250 | 59 |  | Women in their third trimester of pregnancy (>28W) | Secondary | n/a | **Geographical origin** **(pregnant women), Geographical origin (partner)** – Belgian, European, non-European | Indirect | 75% | M |
| Leinander and Olsson (28) | 2017 - 2018 | Sweden | Qualitative | Semi-structured interviews | Not specified (but school setting) | 7 | 0 |  | Female school nurses who had at least 1 year of experience working with newly arrived migrant children | Primary | Access | n/a | Indirect | 90% | H |
| Louka et al. (29) | 2017 - 2018 | Greece & Netherlands | Qualitative | Interviews | Not specified | 61 | 61 |  | Adult asylum seekers and refugees arriving in host countries at least 4 months prior to the study start date | Primary | Access, Awareness | n/a | Direct | 56% | M |
| Lutgehetmann et al. (30) | 2006 - 2007 | Germany | Cross-sectional | Questionnaire based interviews | Hepatitis B | 201 | 162 | 80.60% | Patients attending a university medical centre with chronic hepatitis B, who had been diagnosed within 1 year | Primary | Access, Awareness | n/a | Direct | 67% | M |
| Mansor-Lefebvre et al. (31) | 2013 | France | Cross-sectional | Questionnaire guided interviews | Childhood immunisations: DTP, MMR, Hepatitis B | 450 | 100 |  | Children of homeless families living in Paris | Both | Access, Affordability | **Age, healthcare contact, French-language**  **Difficulties (parent), educational level (parent), monthly household income, health insurance, number of residence changes,** gender, school attendance, other family members in greater Paris area. | Indirect | 100% | H |
| Marek et al. (32) | 2014 - 2015 | Hungary | Cross-sectional | Questionnaire | Not specified | 123 | 123 |  | Asylum seekers who attended didactic educational programme in reception centre | Both | Access, Awareness | Gender, Age, **Geographical origin** (SE Europe, **Africa**, Western Asia, Middle East), Length of stay in Hungary, Current migrant status | Direct | 78% | M |
| McGeown, Moore and Heffernan (33) | 2015 - 2016 | England, UK | Qualitative | Semi-structured face-to-face interviews | Not specified | 300 practices | 0 |  | Staff at London GP practices | Primary | Access, Awareness | n/a | Indirect | 80% | H |
| Mellou et al. (34) | 2017 - 2018 | Greece | Cross-sectional | Evaluation of vaccination campaign | MMR, DTP, polio, pneumococcal disease, Hib, Hepatitis B | 3786 | 3786 |  | Refugees, asylum seekers and newly arrived migrants’ children | Both | Access | Sex, **geographical origin, camp size** | Direct | 80% | H |
| Meynard et al. (35) | 2010 - 2011 | Switzerland | Cross-sectional | Analysis of medical records | MMR, Tetanus, Hepatitis B, HPV | 133 | N/A | | Young people living in Switzerland | Secondary | n/a | Age, sex, **length of stay** | Direct | 63% | M |
| Mikolajczyk et al. (36) | 2004 - 2005 | Germany | Cross-sectional | Analysis of medical records | MMR, Hepatitis B | 1481 | 265 |  | Pre-school children | Secondary | n/a | Gender, **acculturation,** parental education level, sole or joint parenting, apartment size, satisfaction with financial situation, household size | Direct | 88% | H |
| Moller, Kristiansen and Norredam (37) | 2008 - 2012 | Denmark | Cohort | Register-based nationwide cohort study | HPV | 22,848 | 3,264 |  | Adolescent refugee girls under the age of 18 | Secondary | n/a | **Geographical origin, current migrant status, length of stay, income**, age | Direct | 100% | H |
| Moura and Rosario O. Martins (38) | 2013 - 2014 | Portugal | Cross-sectional | Analysis of data from 2014 National Health Survey | Tetanus | 1277 | 1277 |  | Adult migrants | Both | Access, Affordability | Gender, **age**, marital status, education level, employment status, **household income**, degree of urbanisation, **region of residence,** self-perceived health status, region of birth, length of stay, **not having** **citizenship**, **private health insurance**, **contact with GP** | Direct | 89% | H |
| Nakken Norredam & Skovdal (39) | 2015 - 2016 | Denmark | Qualitative | Semi-structured interviews | Childhood vaccination/not specified | 6 | 0 |  | Public health nurses and doctors working at 2 Red Cross asylum centres | Primary | Access | n/a | Direct | 90% | H |
| Nakken et al. (40) | Active cases as of Oct 2015 | Denmark | Cross-sectional | Retrospective database analysis | Not specified | 2126 | 2126 |  | Asylum-seeking children (aged 3 months - 17 years) | Both | Access | **Geographical origin, age, gender** | Direct | 100% | H |
| Napolitano et al. (41) | 2016 – 2018 | Italy | Cross-sectional | Structured questionnaire-guided interviews | HPV | 427 | 427 |  | Immigrants and refugees visiting hospital outpatient care | Primary | Awareness | n/a | Direct | 78% | M |
| Perry et al. (42) | 2014 - 2018 | Wales, UK | Cross-sectional | Database analysis and questionnaire-guided interviews | Childhood immunisations: DTaP/IPV/Hib, MMR, Hib/MenC, Td/IPV, MenACWY | 388 | 388 |  | Accompanied asylum-seeking children aged 5-16 years | Both | Access | **Dispersal site, migrant status** (asylum seeker versus local population) | Direct | 78% | M |
| Poethko-Müller et al. (43) | 2003 - 2006 | Germany | Cross-sectional | National survey | Measles | 14,826 | 3,253 |  | Sample of children living in Germany | Secondary | n/a | **Migration background (parents), migration background (foreign-born vs German-born),** Geographical origin | Direct | 100% | H |
| Redsell et al. (44) | N/A | England, UK | Qualitative | Semi-structured interviews | Childhood immunisations/MMR, others not specified | 22 | 0 |  | Health visitors | Primary | Access | n/a | Indirect | 80% | H |
| Ricco et al. (45) | 2010 - 2012 | Italy | Cross-sectional | Standardised questionnaire and analysis of medical records | Tetanus | 554 | 156 |  | Unskilled (manual labour) construction workers | Both | Awareness, Acceptance, Activation | **Geographical origin** | Indirect | 78% | M |
| Rondy et al. (46) | 2009 | Netherlands | Cross-sectional | Evaluation of vaccination campaign | HPV | 381,869 | 24,643 |  | Girls born between 1993-1996 | Secondary | n/a | **Geographical origin (parents)** | Indirect | 88% | H |
| Salad et al. (47) | 2015 | Netherlands | Qualitative | Semi-structured interviews and group discussion | HPV | 20 | 20 |  | Somali women | Primary | Acceptance | n/a | Direct | 90% | H |
| Santorelli et al. (48) | 2008 – 2012 | England, UK | Cohort | Analysis of medical records | Childhood Immunisations: DTaP/IPV/Hib, MenC, PCV, Hib/MenC, MMR, DTaP/IPV | 6977 | N/A | | Sample of children born in Bradford, UK | Secondary | n/a | **Geographical origin (mother; UK/not UK)** | Indirect | 100% | H |
| Slåttelid Schreiber et al. (49) | 2009 – 2011 | Denmark | Cohort | Register-based nationwide cohort study | HPV | 65,926 | 6,280 |  | Girls born between 1996 and 1997 | Secondary | n/a | Birth cohort, **migration background (descendant/immigrant),** **mother’s disposable income** | Indirect | 100% | H |
| Sim et al. (50) | 2009 | Scotland, UK | Qualitative | Semi-structured interviews | Influenza | 10 | 5 |  | Pregnant Polish women | Primary | Acceptance, Access | n/a | Direct | 90% | H |
| Suppli et al. (51) | 2014 – 2015 | Denmark | Cohort | Nationwide register-based cohort study | HPV, MMR | 9692 | 1386 | 14.3% | Danish girls and their foreign-born parents | Primary | Activation | n/a | Indirect | 91% | H |
| Van der Wal et al. (52) | 2003 | Netherlands | Cross-sectional | Register-based retrospective cross-sectional study | DTP, MMR | 57,382 | 34,305 |  | Children aged between 5 and 12 years, living in Amsterdam | Both | Access, Activation | **Geographical origin** | Direct | 100% | H |
| Van Lier et al. (53) | 2009 | Netherlands | Cross-sectional | Database analysis | Childhood Immunisations: DTaP/IPV/Hib, MMR, MenC | 180, 456 | 37,641 |  | Children of migrants | Secondary | n/a | **Geographical origin, migration background (one/two sided)** | Indirect | 100% | H |
| Vandermeulen et al. (54) | 2003 | Belgium | Cross-sectional | Questionnaire and analysis of medical records | MMR | 3490 | N/A | | Children and adolescents residing in Belgium | Secondary | n/a | **Age, geographical origin (mother)** | Indirect | 100% | H |
| Vilajeliu et al. (55) | 2008 - 2013 | Spain | Cross-sectional | Analysis of medical records and serological studies | Rubella | 22,674 | 9,406 |  | Mothers who had given birth between 2008-2013 | Secondary | n/a | **Geographical origin** | Indirect | 88% | H |
| Vita et al. (56) | 2013 - 2017 | Italy | Cross-sectional | Database analysis | Hexavalent (diphtheria, tetanus, pertussis, polio, Hib, Hepatitis B), MMRV, pneumococcal, MenC, Hepatitis B, DTaP/IPV, polio, HPV | 3941 | 3941 |  | Asylum seekers living in a residential institution | Primary | Access | n/a | Direct | 88% | H |
| Walter et al. (57) | 2013 | Germany | Qualitative | Focus group discussions | Not fully specified but including HPV and influenza | 80 | 72 |  | Young people, migrant mothers and doctors | Primary | Awareness, Acceptance, Activation | n/a | Direct | 80% | H |
| Waxenegger et al. (58) | 2013 - 2015 | Austria | Cross-sectional | Database analysis | Childhood immunisations/not specified | 5,227 | 632 |  | Migrant children living in Austria | Secondary | n/a | **Geographical origin** | Direct | 75% | M |
| Zeitlmann, George and Falkenhorst (59) | 2013 - 2014 | Germany | Project evaluation | Evaluation of new initiative | Polio | 33,874 | 33,874 |  | Asylum seekers | Primary | Access | n/a | Direct | N/A | |
| Bjerke et al. 2021 (60) | 2009-2014 | Norway | Cohort | National Register-based retrospective cohort study | HPV | 177,387 | 18,649 |  | Girls born in Norway | Secondary | n/a | **Geographical origin; parental education level** [unclear which other associations measured] | Direct | 100% | H |
| Deal et al. 2021 (61) | 2020-2021 | UK | Qualitative | In-depth semi-structured interviews | COVID-19 | 32 | 32 |  | Migrants (refugees, asylum seekers, undocumented migrants and migrants with limited leave to remain) | Primary | Acceptance, Affordability, Access, Awareness | n/a | Direct | 80% | H |
| Ganczak et al. 2021 (62) | 2019 | Poland | Qualitative | Semi-structured focus group discussions | Not specified | 22 | 22 |  | Ukrainian migrants living in Poland. Majority had lived in Poland for less than 4 years. | Primary | Access, Acceptance, Awareness | n/a | Direct | 100% | H |
| Jenness et al. 2021(63) | 2000-2016 | Norway | Cross-sectional study | National population register | Measles | 11,334 | 11,334 |  | All children born in Norway from 2000-2016 where both parents originated from Somalia | Secondary | n/a | **Mother’s length of stay, residential area, gender – birth year interaction,** birth year | Direct | 75% | M |
| Moussaoui et al. 2021 (64) | 2017-2018 | France | Cross-sectional | Online questionnaire | Catch-up vaccination/not specified | 216 | N/A |  | GPs involved in the care of migrants | Primary | Access | n/a | Indirect | 75% | M |
| Spadea et al. 2021 (65) | 2009-2014 | Italy | Cohort | Multicentre retrospective birth cohorts | Childhood immunisations: tetanus, measles, MenC | 23,287 | 23,287 |  | Children born in 2009-2014 to foreign women in Rome, Turin and Treviso | Secondary | n/a | **Geographical origin (mother)** | Direct | 73% | M |
| Bell et al. 2020 (66) | Approx. 2018 | UK | Qualitative | Semi-structured interviews | Measles | 42 | 9 migrants; 33 providers |  | Providers involved in vaccination delivery and outbreak management; Romanian and Roma migrants | Primary | Access, Affordability, Awareness, Activation, Acceptance | n/a | Direct | 80% | H |
| Knights et al. 2021 (67) | 2020-2021 | UK | Qualitative | Semi-structured interviews | COVID-19 | 81 | 17 migrants; 64 clinicians |  | Clinical primary care professionals and recently-arrived migrants | Primary | Access, Acceptance, Awareness | n/a | Direct | 100% | H |

**Table S 2. Statistically significant (p<0.05) determinants of under-vaccination in migrants and details, presented by citation, study location and population.**

| Study authors | Location | Population  (type of migrant) | Determinants of under-vaccination identified | | | | | Details of determinants statistically significantly associated (p<0.05) with under-vaccination |
| --- | --- | --- | --- | --- | --- | --- | --- | --- |
|  |  |  | Geographical origin | Sex | Age | Length of residence | Other^1^ |  |
| Amdisen et al. (1) | Denmark | Migrant adolescents |  |  | x |  | x | *Being born 1999-2000; first generation status (implying lower level of acculturation)* |
| Borras et al. (3) | Spain | Migrant children | x |  |  |  | x | *African origin (parental); parents who spent <12 years in education; mothers ≤ 30 years old; families with >3 children.* |
| Bouhamam et al. (4) | France | Migrant children | x |  |  |  | x | *Parents of Eastern European origin; Language difficulties* |
| Brockmann et al. (5) | Germany | Migrant children |  |  | x |  |  | *Older age* |
| Cleary et al. (8) | Ireland | Pregnant women | x |  |  |  |  | *Eastern European origin, African origin, Asian and Middle Eastern origin* |
| De Oliveira Bressane Lima et al. (9) | Netherlands | Migrant adolescents | x |  |  |  |  | *Moroccan or Turkish origin* |
| Fabiani et al. (11) | Italy | Migrants | x |  |  | x | x | *Migrants from high migratory pressure countries in sub-Saharan Africa and Asia; living in Italy <5 years; region of residence (living in Northern Italy)* |
| Fernandez de Casadevante et al. (12) | Denmark | Migrants | x |  |  | x |  | *Being foreign-born versus descendant; lived in Denmark for 6-10 years.* |
| Fozouni et al. (13) | Germany | Refugees | x |  | x |  | x | *Syrian children under 5 years cf Afghan children under 5 years; Age: Syrian children under 5 years old cf those 5 years and older; Recent arrival at camp; Paternal unemployment; Lower parental educational level* |
| Freund et al. (14) | France | Pregnant women | x |  |  |  |  | *Sub-Saharan origin; North African origin; Asian origin* |
| Hansen et al. (20) | Norway | Migrant adolescents | x |  |  |  |  | *Parents with African, Central and South American or Old EU/Western origin* |
| Hertzum-Larsen et al. (22) | Denmark | Migrant adolescents | x |  | x |  |  | *Geographical origin (Mid and Eastern Asia, North Africa and Western Asia, Eastern Europe, South and Central America, Sub-Saharan Africa, Western countries, cf Denmark in birth cohort 1996-2000 and Mid and Eastern Asia, Eastern Europe, South and Central America, Sub-Saharan Africa, Western countries (all regions except for North Africa and Western Asia) in birth cohort 2012-2015); being born 1996 - 2000* |
| Jimenez-Garcia et al. (24) | Spain | Migrants |  |  | x |  |  | *Migrants aged >=65 (cf native Spanish population) were less likely to have received the influenza vaccine (aOR 0.85 (95%CI 0.35-2.06)* |
| Jimenez-Garcia et al. (25) | Spain | Migrants |  | x | x |  | x | *Compared to native population, migrants had significantly (p<0.05) lower vaccination coverage by age, sex, comorbidity, and risk group.* |
| Laenen et al. (27) | Belgium | Pregnant women | x |  |  |  |  | *Non-European Origin* |
| Mansor-Lefebvre et al. (31) | France | Migrant children | x |  | x |  | x | *DTP-IPV: children aged 6-9; no contact with health system in previous year; children born outside France; children who had parent with French language difficulties; changed residence at least twice in previous year.  MMR: children born outside France; children who had parent with French language difficulties, changed residence at least twice in previous year; no contact with health system in previous year; no health insurance; parent had primary or middle school education compared to none. HepB: children aged 6-9; no contact with health system in previous year; children born outside France; higher household income.* |
| Marek et al. (32) | Hungary | Asylum seekers | x |  |  |  |  | *African origin* |
| Mellou et al. | Netherlands and Greece | Refugees & asylum seekers, newly arrived migrants | x |  |  |  | x | *Syrian and Iraqi children had lower vaccination coverage than Afghan children (p<=0.001); smaller camps had lower MMR coverage compared to larger camps (p=0.016) – trend also observed for other vaccines without statistical significance* |
| Meynard et al. (35) | Switzerland | Migrants |  |  |  | x |  | *Recently immigrated (<2 years ago)* |
| Mikolajczyk et al. (36) | Germany | Migrants |  |  |  |  | x | *Lower level of acculturation* |
| Moller, Kristiansen and Norredam (37) | Denmark | Refugees & asylum seekers | x |  |  | x | x | *Having had a residence permit ≥ 5 years; being a refugee; lower household income; Middle East, African, Eastern European, Afghan and former Yugoslavian origin* |
| Moura and Rosario O. Martins (38) | Portugal | Migrants |  |  | x |  | x | *Age >65 years old; higher household income; Portuguese citizenship; no private health insurance; last contact with GP >=12 months ago; region of residence (live in Lisboa, Algarve or RA da Madeira)* |
| Nakken et al. (40) | Denmark | Asylum seeking children | x | x | x |  |  | *Afghan, Eritrean, Iranian, Russian, Iraqi origin; being female; older children aged 12-17.* |
| Perry et al. (42) | United Kingdom | Asylum seeking Children |  |  |  |  | x | *Asylum dispersal site; migrant status (asylum seeker c.f. local population)* |
| Poethko-Muller et al. (43) | Germany | Migrant children |  |  |  |  | x | *First generation migrant (foreign-born); one-sided migration background* |
| Ricco et al. (45) | Italy | Migrants | x |  |  |  |  | *Eastern Mediterranean Region (EMR) origin* |
| Rondy et al. (46) | Netherlands | Migrant adolescents | x |  |  |  |  | *Turkish or Moroccan origin* |
| Santorelli et al. (48) | United Kingdom | Migrant children | x |  |  |  |  | *Children of foreign-born White British mothers (c.f. UK-born White British women); [children of foreign-born Pakistani mothers were more likely to be fully immunised than Pakistani children whose mothers were UK-born]* |
| Slåttelid Schreiber et al. (49) | Denmark | Migrant adolescents |  |  |  |  | x | *First generation migrant (foreign-born); higher household income; higher maternal education [data not shown]* |
| Van der Wal et al. (52) | Netherlands | Migrant children | x |  |  |  |  | *Moroccan, Turkish or Surinamese origin* |
| Van Lier et al. (53) | Netherlands | Migrant children | x |  |  |  | x | *One- or two-sided migration background; other Western and non-Western origin* |
| Vandermeulen et al. (54) | Netherlands | Migrant children | x |  |  |  |  | *Adolescents with mothers of non-European origin; primary school children with mothers of European origin* |
| Vilajeliu et al. (55) | Spain | Pregnant women | x |  |  |  |  | *Asian, African, or American origin* |
| Waxenegger et al. (58) | Austria | Migrant children | x |  |  |  |  | *Non-European origin* |
| Lutgehetmann et al. 2010 (30) | Germany | Regular migrants |  |  |  |  | x | *Poor host country language skills* |
| Bjerke et al 2021 (60) | Norway | Migrant children | x |  |  |  | x | *Girls with parents from Western Europe, Central and Eastern Europe, Sub-Saharan Africa, America, or Oceania; Higher parental education* |
| Jenness et al 2021 (63) | Norway | Migrant children | x | x |  | x |  | *Children of mothers of Somali origin; mother’s residence in Norway ≥6 years; children born in Oslo and Akershus county; boys born in more recent years* |
| Spadea et al 2021 (65) | Italy | Migrant children | x |  |  |  |  | *Parents of Asian origin* |

^1^ Other category includes: Being less acculturated, being a refugee/asylum seeker, income, not having accessed healthcare/GP in past 12 months, not having private health insurance, having frequent residence changes, specific region of residence, specific asylum dispersal site, living in a smaller refugee camp, not having citizenship, having a comorbidity, being in an influenza risk group, younger maternal age, parental education level, parents’ difficulties speaking host country language, parents unemployed, one/both parents born overseas, first generation migrant children, larger family size.

Notes: Five studies whose data were not included for the secondary outcome due to lack of statistical test showed lower rates of coverage in African migrants (Blackwell; Borras); higher indication (need for) HBV vaccine in Eastern European asylum seekers and lower vaccination coverage in undocumented children of non-Brazilian nationality compared to undocumented children of Brazilian nationality.

- Blackwell et al. (68): Positive BCG vaccination coverage by origin: Middle East (189/ 211, 89.6%), Eastern Europe (93/120, 77.5%) Asia (17/22, 77.3%), Africa (23/42, 54.8%).
- Serre-Delcor et al. (69): HBV vaccine indicated in 86.7% of Eastern European asylum seekers
- Klok-Nentjes et al. (26): Undocumented children of non-Brazilian nationality had lower vaccination coverage cf Brazilian nationality
- Borras et al. (3) found coverage of the 4:4:4:3:1 schedule was lowest in migrants of African origin (South American origin (83.3%, Oceania (75%), Europe (68.4%), Africa (64%)).
- Parellada et al. (70): Increasing number of older undocumented migrant children do not get vaccinated or get them later than recommended

**Table S 3. Example search terms: keywords (consistent across all databases).**

| **Theme** | **Keywords** |
| --- | --- |
| Migrants | migrant* OR immigrant* OR emigrant* OR foreign-born OR “foreign born” OR foreign-origin OR “foreign origin” OR foreign* OR “asylum seek*” OR asylum-seek* OR asylee* OR refugee* non-citizen* OR citizenship OR nationality OR undocumented OR non-resident* OR expat* OR newcomer* OR new-comer |
| Vaccination | vaccin* or immunis* or immuniz* or MMR or MMRV or BCG |
| Determinants | (uptake or demand or coverage or utiliz* or utilis*) OR  (barrier* or enabl* or facilitat* or motivat* or obstacle* or determinant* or factor* or reason* or challenge*) OR  (accept* or comply or complian* or adher* or readiness or intent* or willing*) OR  (avoid* or refus* or hesita* or renounc* or reject* or deny or deni* or delay*) OR  (confiden* or trust* or fear* or wary or wariness or doubt* or sceptic* or concern* or complacen*) OR  (attitude* or perception* or perspective* or view* or belief*) OR  (practice* or behavio?r*) OR (decision or decision-making) OR  (aware* or knowledge* or inform* or understand*) OR  (access* or cost* or afford* or navigat* or availab*) |
| Year | Limit [no.] to yr="2000 – 2021” |

**Table S 4. Example search terms: MeSH terms (varied for each database; example below used for MEDLINE).**

| **Theme** | **MeSH term** |
| --- | --- |
| Migrant | Exp Transients and Migrants/ Exp Emigrants and Immigrants/  Exp Refugees/ |
| Vaccination | Exp Vaccination/  Exp Mass Vaccination/  Exp Anti-Vaccination Movement/  Exp Vaccination Refusal/  Exp Vaccination Coverage/  Exp Immunization/ |
| Determinants | Exp Health Knowledge, Attitudes, Practice/ Exp Health Services Accessibility/ |

**Table S 5. Barriers identified in included studies, by domain, level (patient, provider, system) and citation.**

| Domain | Barriers | Level | | |
| --- | --- | --- | --- | --- |
|  |  | **Patient** | **Provider** | **System** |
| Access | Do not speak the language or not fluent; no interpreter (4, 6, 16, 18, 23, 30, 31, 33, 44, 47, 59, 61, 66, 71) | X | X |  |
|  | Low literacy (23, 44, 66) | X |  |  |
|  | Lack of tailored or translated information or inappropriate format (4, 6, 16, 21, 31, 33, 44, 47, 57, 61, 62, 66) |  |  | X |
|  | Limited appointment time (66) |  | X |  |
|  | Insecure housing; frequent change of address (15, 28, 31) | X |  |  |
|  | Stage of migration (e.g. transit versus destination) (29, 32, 34) | X |  |  |
|  | Lack of opportunity or service provision, including due to government policy (29, 32, 72) |  | X | X |
|  | Missed opportunities to vaccinate (38) |  | X |  |
|  | Means of determining vaccination history too resource intensive (28, 59) |  | X |  |
|  | Resource and capacity constraints (23, 34, 42, 59) |  | X |  |
|  | Experience or fear of discrimination, distrust of authorities/health system; sense of abandonment (15, 23, 61, 62, 66, 67) | X | X | X |
|  | Variability in local procedures, resource allocation and coordination (23, 34, 42, 59, 64) |  | X | X |
|  | Practical and legal barriers to accessing health services (15, 23, 67) |  |  | X |
|  | Access points, e.g. unfamiliar or inaccessible settings; delivering routine immunisations through schools (23, 61) | X |  | X |
|  | Providers lack awareness of policy and guidelines, e.g. entitlements, catch-up vaccination for migrants or other countries' vaccination schedules (15, 29, 33, 52, 62, 64) |  | X |  |
| Affordability (financial) | Cost – direct (6, 12, 29, 66) | X | X | X |
|  | Cost – indirect (e.g. travel costs) (61) | X |  |  |
| Affordability (non-financial) | Competing priorities; attending vaccination appointment or health check-up is low priority; pre-booked appointments create inflexibility (15, 61, 66) | X |  |  |
| Awareness | Lack knowledge of disease or link between the disease and vaccine (6, 18, 41, 47, 67) | X |  |  |
|  | Lack knowledge of own medical history or vaccination status (19, 32) | X |  |  |
|  | Lack knowledge of entitlement to vaccination/primary care (23, 32) | X |  |  |
|  | Low health literacy/knowledge of need for vaccination or boosters (6, 10, 17, 23, 29, 30, 32, 41, 45, 57, 62, 66) | X |  |  |
|  | Unfamiliar with immunisation schedule or need for boosters (33, 45, 66) | X |  |  |
|  | Unaware that a vaccine is available (41) | X |  |  |
|  | Cannot locate credible or trusted information about the vaccine/vaccination (61, 62, 66, 67) | X |  | X |
| Acceptance | Worries about vaccination, vaccine safety, side effects (particularly of specific vaccines e.g. HPV), fear of autism, activities of anti-vax groups, etc. (10, 15-18, 23, 45, 50, 57, 61, 66) | X |  |  |
|  | Concerns around new vaccines and whether they are safe (21, 61) | X |  |  |
|  | Refused vaccination for religious/personal reasons (45, 47) | X |  |  |
|  | Alienation and disempowerment; distrust of the health system; distrust the motives behind vaccination (e.g. microchip, sterilisation); fear of being questioned about immigration status (15, 47, 61, 66, 67) | X |  | X |
|  | Misinformation from internet, social media, family, friends and unofficial sources due to lack of tailored information sources they can access (16, 50, 67) | X |  |  |
|  | Negative social norms around vaccination in country of origin - e.g. unfashionable, vaccination in pregnancy not recommended (16, 50) | X |  |  |
|  | Cultural acceptability, norms, and stigma around specific vaccines (18, 39, 47, 57) | X |  |  |
|  | Low perceived value or importance of vaccination or disease risk (10, 18, 29, 57, 61, 62, 66, 67) | X |  |  |
|  | Lack of information (10, 50, 61, 62) | X |  |  |
|  | Vaccination not provider-recommended (10) |  | X |  |
| Activation | Lack of practical support, exchanges and recommendations from health care professionals around vaccination, when they were desired (57) | X | X |  |
|  | Blanket approaches (e.g. vaccination reminders sent via letter/text message) not suitable for transient migrant populations, e.g. Roma (66) |  | X |  |
| Other | Lack vaccine documentation/record, e.g. vaccination card (19, 28, 32, 59) |  |  | X |

**Table S 6. Facilitators identified in included studies, by domain, level (patient, provider, system) and citation.**

| Domain | Facilitator |  | Level |  |
| --- | --- | --- | --- | --- |
|  |  | **Patient** | **Provider** | **System** |
| Access | Attending school (31) | X |  |  |
|  | Social integration; engagement with healthcare system (15, 23, 31, 38, 57) | X |  | X |
|  | Having citizenship (38) | X |  | X |
|  | Local coordination (approach; vaccine supply); partnership working (5, 34, 66) |  | X | X |
|  | Larger camps – organisation of services (34) |  | X | X |
|  | Culturally competent care; migrant-sensitive services and policies (7, 15, 28, 44, 56) |  | X | X |
|  | Trust in the provider/system/State (15, 62) | X |  |  |
|  | Promoting registration with primary care for migrants (61) |  |  | X |
|  | Alternative access points (56, 61) | X | X | X |
|  | Working with communities to co-develop tailored resources (61, 67) | X |  |  |
|  | Access to translated information (44) | X |  |  |
|  | Ease of booking appointments (21, 62) | X | X |  |
|  | Policy to vaccinate in absence of vaccination card/record (56) |  |  | X |
| Acceptance | Perceived importance/benefit of vaccination (15, 17, 18, 21, 71) | X |  |  |
|  | Positive religious beliefs about vaccination (21) | X |  |  |
|  | Normalisation of vaccination (15, 21, 23) | X | X |  |
|  | Perception that vaccines are high quality (16, 62) | X |  |  |
|  | Culturally/religiously tailored health messaging (21, 47) | X | X | X |
|  | Trusted information source (41, 50) | X |  |  |
|  | Provider recommendation (10, 17, 41, 45, 50, 57) | X | X |  |
|  | Adequate and credible information about vaccination (18) | X |  | X |
| Affordability (financial) | Free vaccination; no associated costs (12, 16, 23, 61) | X |  | X |
|  | Having insurance coverage (31, 38) | X |  | X |
| Affordability (non-financial) | Convenience of access, e.g. walk-in clinics rather than pre-booked appointments; flexible appointments (21, 66) |  | X | X |
| Awareness | Health educational programmes (32) |  | X | X |
|  | Aware of benefits of vaccination (17) | X |  |  |
| Activation | On-arrival health screening and vaccination for asylum seekers (40) |  | X | X |
|  | Mandatory vaccination in the workplace (45) |  | X | X |
|  | Provider recommendation (45) |  | X |  |
|  | Mass vaccination campaigns at camps (34) |  | X |  |
|  | Culturally tailored, appropriate, and flexible programmes (34) |  | X | X |
|  | Activities to increase knowledge and awareness of vaccination (29) |  | X |  |
|  | Face-to-face communication; outreach activities (23) | X | X |  |
|  | Personalised reminders (51, 66) |  | X |  |
|  | Community champions and advocates (66) | X |  | X |
